# CovidEnvelope: A Fast Automated Approach to Diagnose COVID-19 from Cough Signals

**DOI:** 10.1101/2021.04.16.21255630

**Authors:** Md Zakir Hossain, Md. Bashir Uddin, Khandaker Asif Ahmed

**Author notes:** {, }.

## Abstract

The COVID-19 pandemic has a devastating impact on the health and well-being of global population. Cough audio signals classification showed potential as a screening approach for diagnosing people, infected with COVID-19. Recent approaches need costly deep learning algorithms or sophisticated methods to extract informative features from cough audio signals. In this paper, we propose a low-cost envelope approach, called CovidEnvelope, which can classify COVID-19 positive and negative cases from raw data by avoiding above disadvantages. This automated approach can pre-process cough audio signals by filter-out background noises, generate an envelope around the audio signal, and finally provide outcomes by computing area enclosed by the envelope. It has been seen that reliable datasets are also important for achieving high performance. Our approach proves that human verbal confirmation is not a reliable source of information. Finally, the approach reaches highest sensitivity, specificity, accuracy, and AUC of 0.92, 0.87, 0.89, and 0.89 respectively. The automatic approach only takes 1.8 to 3.9 minutes to compute these performances. Overall, this approach is fast and sensitive to diagnose the people living with COVID-19, regardless of having COVID-19 related symptoms or not, and thus have vast applicability in human well-being by designing HCI devices incorporating this approach.

## 1 Introduction

COVID-19 is a respiratory disease caused by SARS-CoV-2 virus – a novel Coronavirus of family Coronaviridae. Coronaviruses of this family, especially viruses of genus Betacoronavirus (e.g. Middle East Respiratory Syndrome Coronavirus, aka MERS-CoV, Severe Acute Respiratory Syndrome Coronavirus, aka SARS-CoV etc) are highly pathogens of respiratory tract diseases and their characteristics of highly variable genetic diversity and diverse host adaptability make them deadly and devastating around the world [16]. Current COVID-19 pandemic and its widespread infection and mortality rate has made SARS-Cov2 virus a hot topic for diverse research communities. Beside developing vaccines to cure COVID-19, substantial effort is being made to develop tools to diagnose COVID-19.

COVID-19 diagnoses tools can be widely divided into two categories. Firstly, biological tools, which are involved in viral nucleic acid-based detection (e.g. RT-PCR, LAMP etc.) and protein-based detection (e.g. rapid antigen-based detection and serological tests). Among the available tools, RT-PCR based nucleic acid detection is being considered as “gold-standard” for COVID-19 diagnosis because of its high specificity, sensitivity, and ability to detect at initial stage of infection [7]. Due to substantial similarity with other sister species of coronavirus, two-targeted multiplex RT-PCR is adopted to detect SARS-CoV2 virus, where ‘first target’ broadly detects presence of any members of coronavirus, and the ‘later target’ further narrow down to SARS-CoV2. Although this technique is rapid and highly reproducible, it is labour-extensive, time-consuming, and need molecular biology expertise with sophisticated laboratory facilities to do certain steps [18]. Different modifications of RT-PCR techniques have been proposed, like one-step Loop-mediated iso-thermal amplification reaction (LAMP), Microarray-based methods, bar-coded bead assays [15], which need sophisticated instruments and are not as sensitive as RT-PCR. On the other hand, protein-based detection tests are simple and fast alternatives, utilises host immune response to viral antigens. Currently, numerous serological tests (e.g. Enzyme-linked immunosorbent assay-ELISA, Indirect immunofluorescence- IIFT) are under development for COVID-19 diagnosis with variable specificity. Even though host antibodies, generated in response to viral infection, can be useful tool for COVID-19 diagnosis, there is a potential chance of producing similar antibodies (cross-reactive antibody response), in response to other coronaviruses – resulting in false positive detection [9]. Besides, antibody-based serological tests are also prone to viral lag-period of 4 to 7 days, where they does not show any responses, and also show poor response up to 6 days of infection, which is alarming for public health [5].

Beside biological tests, several clinical feature-based tests are being proposed and analysed. The initial mild symptoms of the COVID-19 include cough, fever, fatigue, followed by headache, dyspnea, myalgia, and gastrointestinal complications with nausea and watery diarrhoea [17], which are being considered during these tests. Severe COVID-19 infection manifested by pneumonia with acute respiratory distress syndrome, severe cough, and infiltrates on chest image. Based on the features, fever, cough, and dyspnea are considered as potential indicator of suspicious COVID-19, and numerous machine-learning (ML) algorithms are being developed as a pre-screening diagnostic tool for COVID-19 detection.

Different ML methods have been used to diagnose various diseases [6, 10] and similar principle has been adopted for diagnosing COVID-19 from chest computed tomography (CT scan) images in [14]. On the other hand, audio signals have been successfully utilised to diagnose various respiratory conditions [12]. Cough detection from the audio signals is a very important and promising process to detect pathology severity of the people, infected with COVID-19. The audio-based screening tool could be implemented in residential environments to track individuals who are suffering from COVID-19 as a subsystem of remote health monitoring systems.

ML has been found as a useful method to design the audio-based screening tool to diagnose coughs [2]. Convolutional Neural Network (CNN) and Recurrent Neural Network (RNN) models were used to detect cough sounds by varying hyper-parameter values manually in [1]. A real-time cough detection method was designed, by combining Gaussian Mixture model and Universal Background model [13], that requires four steps to complete the process: sound pre-processing, segmentation, feature / event extraction, and cough prediction. Wavelet decomposition and statistical parameters were used to detect pneumonia cough [14]. Logistic regression was considered to detect tuberculosis cough from short-term spectral features [3]. Further, Brown et al. [4] used large-scale crowdsourced dataset of respiratory sounds and extracted various features before detecting COVID-19 cough using logistic regression, gradient boosting trees (GBT) and support vector machines (SVM). Laguarta et al. [8] considered CNN for diagnosing COVID-19 cough from extracted features. In this paper, we propose a low-cost envelope approach that does not require any prepossessing steps and extracting features from raw cough signals. In addition, it is a computationally inexpensive method and easy to develop HCI device for healthcare and wellbeing of the communal health. As a HCI response to pandemic, such real-time cough-screening tool will be helpful to pre-screen and later-on diagnose COVID-19 patients and reduce COVID-19 prevalence around the community.

## 2 Methods

We collected cough audio signals from a reliable, publicly available data repository, and designed an automatic approach, which is capable of diagnosing COVID-19 from the raw cough signals.

### 2.1 Cough Dataset

There is insufficient dataset available regarding COVID-19 cough sounds. Among the available datasets, some lack proper annotation and corresponding confirmation. The most reliable and publicly available dataset, used in current study, was collected by the Medina Medical Group4 in Russia.

The dataset contains a total number of readable sound data of 1322, collected in Oct.-Nov.2020 as mentioned in Table 1. It has two types of records – *Verbal Positives*: where subjects confirmed their COVID-19 presence verbally, *Verbal Negatives*: where subjects confirmed their COVID-19 absence verbally. Further the verbal confirmations were verified using laboratory-based PCR tests, generating two types – *Verified Positives*: where the records had been further confirmed to the presence of COVID-19 by laboratory-based PCR tests and *Verified Negatives*: where the records confirmed for the absence of COVID-19 by PCR tests. Verified positives dataset further divisible into two specific groups – *Symptomatic*: where the subjects exhibit COVID-19 specific symptoms and *Asymptomatic*: where the subjects did not show any COVID-19 specific symptoms. In this study, regardless of COVID-19 presence or absence, we formed another type of dataset, namely *‘Matched’* : where verbal confirmation data matched with the laboratory confirmations. This study found 819 *‘Matched’* records, where the number of *‘Matched Positives’* (*i*.*e. Verbal Positives = Verified Positives*) and *‘Matched Negatives’* (*i*.*e. Verbal Negatives = Verified Negatives*) were 381 and 438 respectively. The *‘Matched Positive’* records are further divided into *‘MatchedSymp’* and *‘MatchedAsymp’* based on the observed COVID-19 related symptoms. There are 201 and 108 records who did not show (*‘MatchedAsymp’*) and showed (*‘MatchedSymp’*) COVID-19 symptoms, respectively.

**Table 1.**
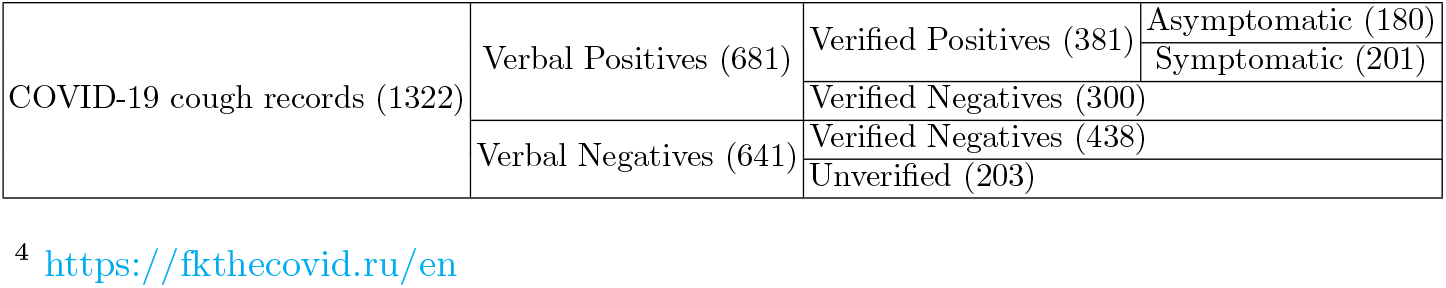
Data specifications with the number of records in parentheses

### 2.2 CovidEnvelope Approach

We designed an envelope approach for computing area of cough sounds which take raw cough audio signals as input and provide outcomes as COVID-19 positive or negative from the computed resultant area. Correct cough-based audio signals were selected from the raw audio signals and then, filtering was performed to get rid of background noises. A “signal envelope” was generated over the filtered audio signal, and the envelope-enclosed resultant area was calculated. Based on the resultant area, decision was made to identify COVID-19 positive or negative. Each step of this automatic approach is described accordingly and a resultant signal is illustrated in Figure 1.

**Fig. 1.**
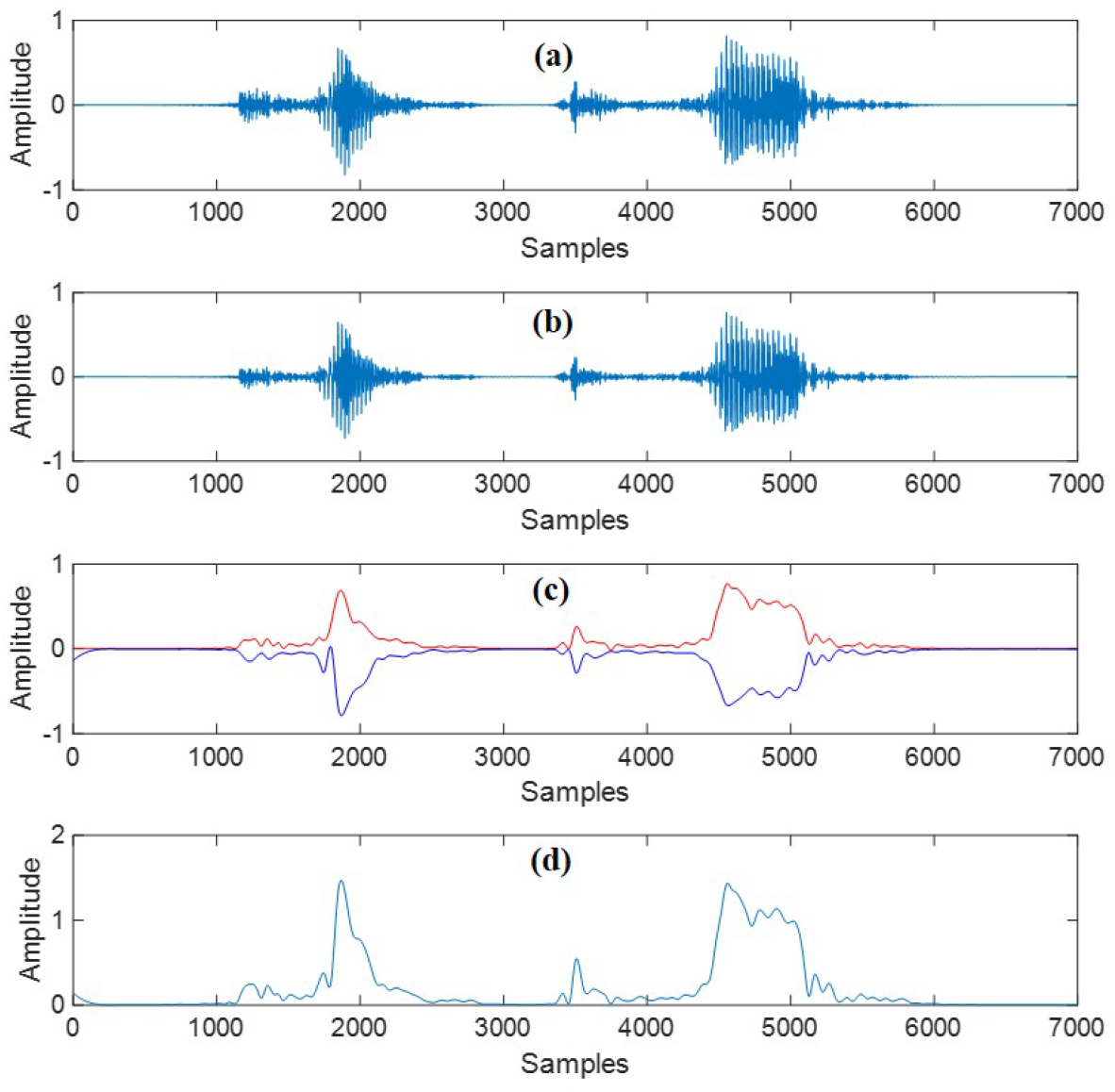
Generating envelope from cough audio signals.

Audio signal, as depicted in Figure 1(a), was taken as input to the algorithm. In addition to the original audio signal, the recording may often contain additional high frequency noisy signals. Correct audio signal was selected by comparing the sum of variances of the recorded signals, and then filtered using a three-point moving average filter to remove random fluctuations between samples of the audio signal as illustrated in Figure 1(b). An envelope of the filtered audio signal was generated as shown in Figure 1(c), where the upper and lower boundaries of the audio signal are shown by the red and blue lines, respectively. The resultant signal was generated by applying equation 1.

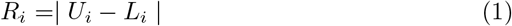

where *R*_*i*_, *U*_*i*_, and *L*_*i*_ are the amplitudes of the resultant, upper, and lower boundaries for the *i*_*th*_ sample. A sample resultant signal is depicted in Figure 1(d). The absolute difference between the upper and lower boundaries was taken to produce positive amplitude enclosed by the signal envelope. The area enclosed by the envelope was calculated by summing up the sample amplitudes of the resultant signal by applying equation 2.

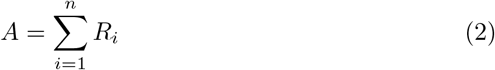

where *‘A’* and *‘n’* are the area enclosed by the envelope and number of samples in the resultant signal, respectively.

## 3 Results and Discussion

We executed our *CovidEnvelope Approach* in an Intel (R) Core (TM) i5-1035G4 CPU@1.10GHz 1.50GHz, 8.00 GB of RAM, Operating system 64-bit computer using MATLAB R2012b and tested with varying thresholds to diagnose COVID-19 positive coughs. Selecting threshold values play a vital role before computing the overall performance. We computed various evaluation matrices against three thresholds for determining optimised threshold values as shown in Figure 2.

**Fig. 2.**
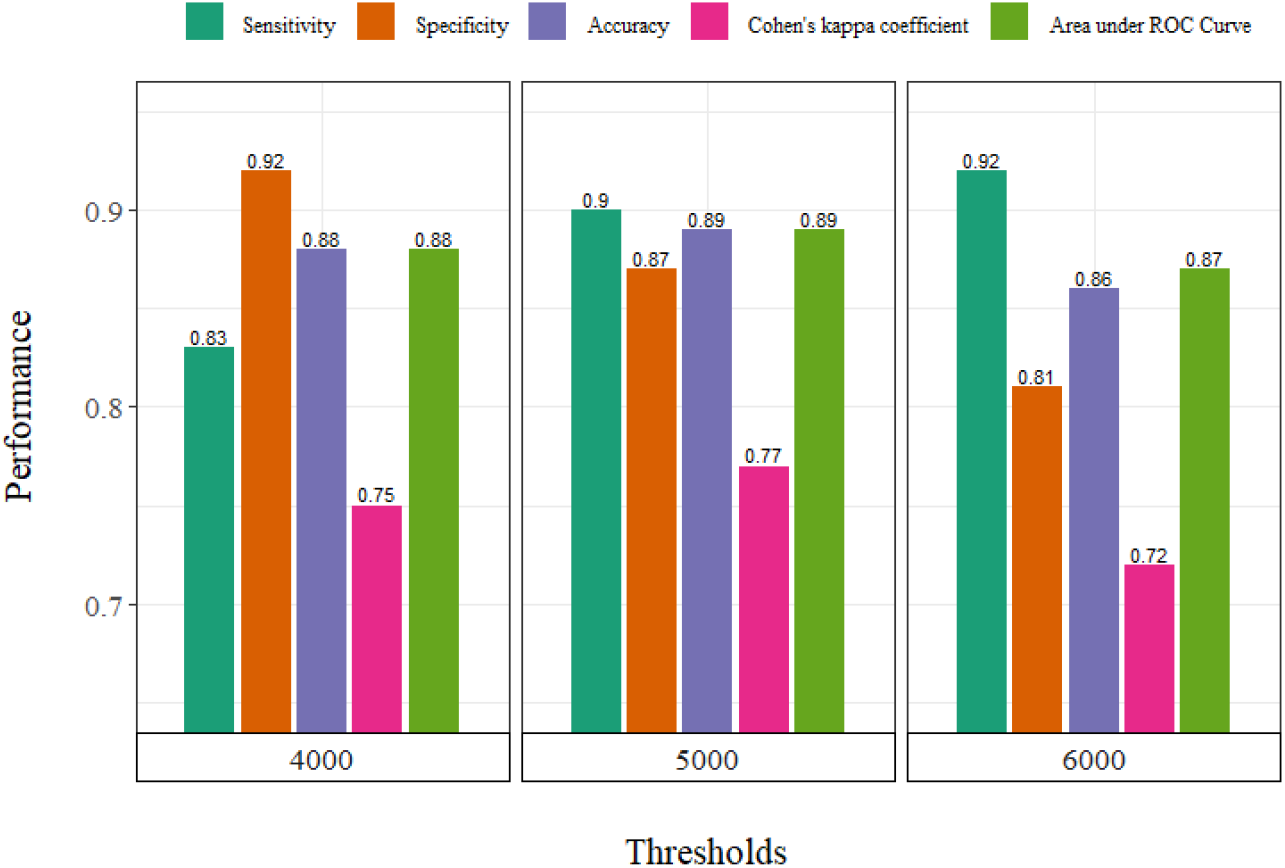
Determination of the optimum threshold value.

For threshold 6,000, sensitivity (0.92) was found highest but other matrices were found lowest compared to other two cases. For threshold 4,000, specificity (0.92) was found highest but other matrices were found lowest compared to threshold 5,000. On the other-hand, for threshold 5,000, accuracy, Cohen’s kappa coefficient (k), and area under ROC curve (AUC) were found highest compared to other two thresholds, which are 0.89, 0.77, and 0.89 respectively. Sensitivity (0.90) and specificity (0.87) are also found reasonable for threshold 5,000. It is worthwhile to note that the accuracy and AUC of the *CovidEnvelope* approach were calculated from the sensitivity and specificity. Thus, we selected threshold 5,000 for further analyses.

We analysed five conditions as explained in Table 2, namely *‘Verbal’, ‘Verified’, ‘Matched’, ‘MatchedAsymp’*, and *‘MatchedSymp’*. The lowest performance (Accuracy = 0.67, AUC = 0.65) was found for *‘Verbal’* condition where the dataset consisted of verbal confirmation verified or unverified with PCR tests. Verbal confirmations are not often correct, prone to miscommunications or fraudulence among participants, and it could be a reason for the lowest performance. When *‘Verified’* condition was considered for COVID-19 dataset, our approach performed better (Accuracy = 0.80, AUC = 0.82) than the *‘Verbal’* condition. It is the reliable records where COVID-19 cases were confirmed by laboratory tests. Performance was improved when verbal confirmation was matched with verified condition (*‘Matched’* condition). Sensitivity, specificity, k, accuracy, and AUC were observed 0.90, 0.87, 0.77, 0.89, and 0.89, respectively for the *‘Matched’* condition. For matched asymptomatic (i.e. *‘MatchedAsymp’*) and symptomatic (*‘MatchedSymp’*) conditions, the *CovidEnvelope* approach reaches up to accuracies of 0.89 and 0.88 respectively, which is very similar to the *‘Matched’* condition. Our approach takes only 1.8 to 3.9 minutes for diagnosing COVID-19 cases depending on an applied condition.

**Table 2.**
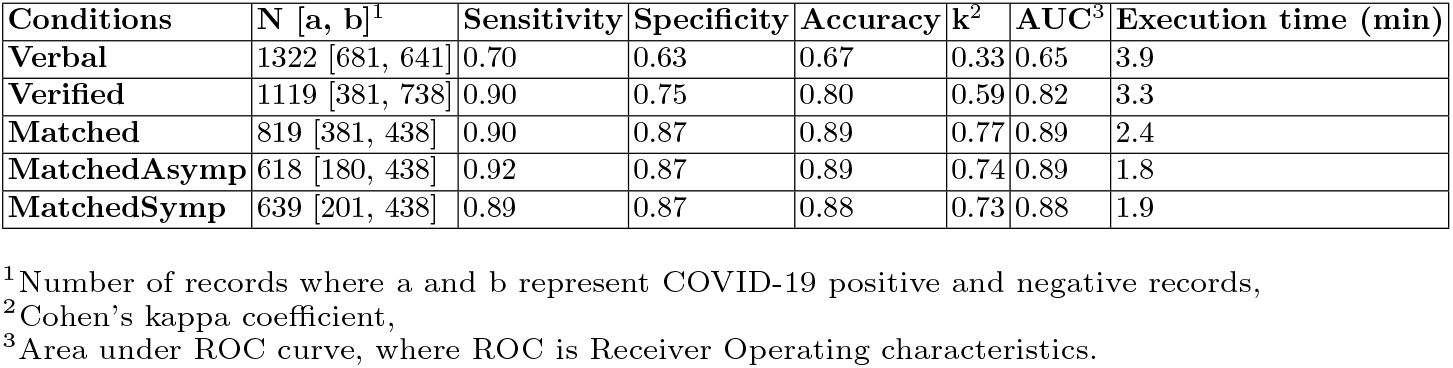
Performance evaluation of the designed algorithm with different conditions

A t-test was performed on computed resultant mean areas for measuring significance tests considering each conditions separately. The computed mean areas are illustrated in Figure 3. For *‘Verbal’* condition, mean areas of COVID-19 positive cases were found significantly different (p = 0.004) than COVID-19 negative cases. For other conditions (*‘Verified’, ‘Matched’, ‘MatchedAsymp’*, and *‘MatchedSymp’*), mean areas of COVID-19 positive cases were found highly significantly different (p*<*0.0001) than COVID-19 negative cases as shown in Figure 3(a). The results indicate that *‘*Verbal*’* confirmation is less reliable than verified and matched conditions for designing an efficient and automatic COVID-19 diagnosis tool. In addition, the mean areas of asymptomatic COVID-19 positive and symptomatic COVID-19 positive cases are illustrated in Figure 3(b) and the mean areas of these cases were not statistically significantly different (p = 0.26). The results alternatively indicate that the performance of our approach is independent of the symptoms of COVID-19.

**Fig. 3.**
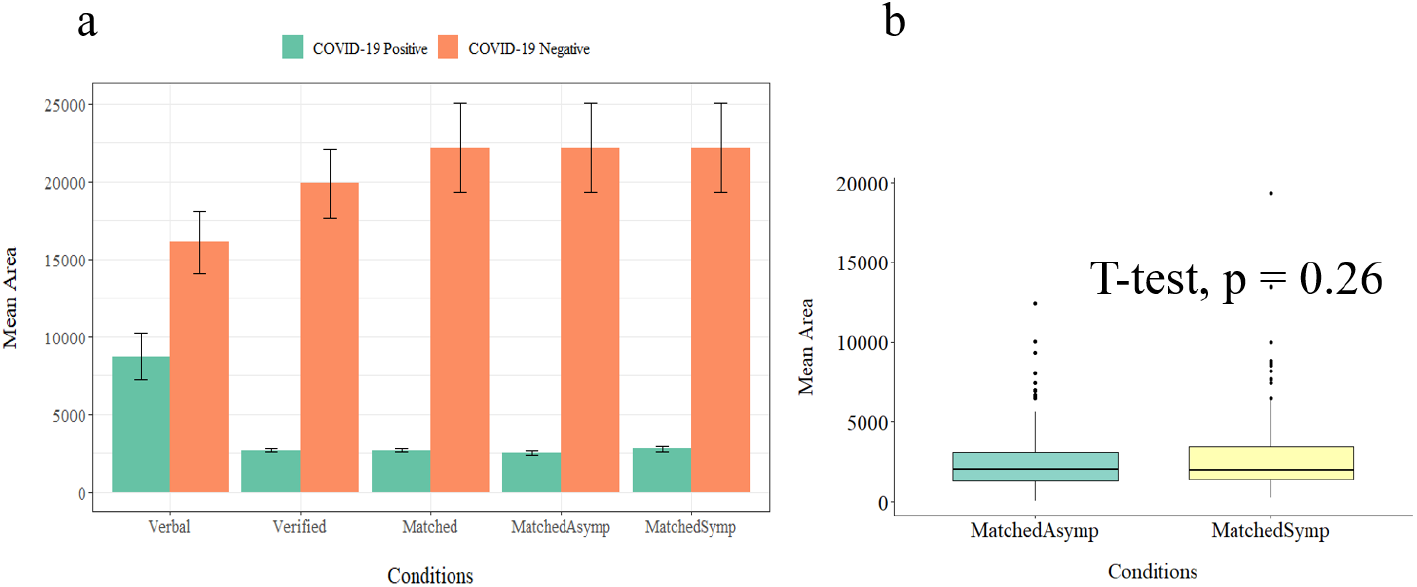
Mean areas computed by *CovidEnvelope Approach*.

The main challenge of this study was to identify trustworthy and reliable datasets. There are several publicly available datasets [4, 12], which were verbally confirmed, but can introduce biases in our approach, as we have seen anomaly in verified and unverified dataset. Till date, MIT Open Voice dataset is the largest COVID-19 cough dataset, comprising 5,320 subjects, resulted in highest sensitivity and specificity of 0.98 and 0.94 respectively, but unavailability of this dataset hinders us to utilise it [8]. Moreover, current dataset does not mention about the level of severity of COVID-19 patients. Using well-documented dataset to validate our approach will enhance both sensitivity and specificity.

Further, our study has shown a compact ML approach, which only needs raw audio signals, do not need to extract any features, and can be recorded easily using available devices like cell phones. Unlike existing approaches, it is computationally inexpensive and requires only less than 4 minutes to screen. Our approach can distinguish between COVID-19 positive and negative coughs, regardless the patients show symptoms or not. Our study will help human well-being by developing tools for hand-hold devices, such as mobile phones, smart watches etc, which would enable diagnosing COVID-19 immediately and improving human-device interaction in pandemic.

The *CovidEnvelope* approach can also be utilised to study other respiratory tract diseases in human, such as tuberculosis, asthma, pneumonia etc. For audio-signal based studies, like classification and surveillance of animals based on audio signals [11], this enveloped approach will add a new horizon. As an extension of current study and potential application in HCI-based healthcare and public health sectors, real-time low-cost software is possible to design in near future. Such a sophisticated, end-to-end encrypted application will need considerable amount of verified COVID-19 records to validate our approach.

## 4 Conclusion

We developed a fast, low-cost, and reliable COVID-19 cough detection approach, which can diagnose COVID-19 with the highest accuracy, AUC, sensitivity, and specificity of 0.89, 0.89, 0.92, and 0.87, respectively. We proved that COVID-19 positive and negative coughs are significantly different in terms of area enclosed by envelope and highly effective regardless of symptomatic or asymptomatic cases. Further, verbal confirmation is not a reliable source of information. Due to the lack of reliable datasets, we are unable to design a pre-trained model. Our future work will focus on collecting more well-documented datasets, especially cough sounds resulting from other pulmonary diseases, by collaborating with relevant authorities, and develop a pre-trained model and a reliable HCI-based mobile application for screening COVID-19 within short duration of time.

## Data Availability

Data is collected from Medina Medical Group, available online for research community.

https://fkthecovid.ru/en

## Acknowledgment

The authors would like to thank *Medina Medical Group* to make the dataset available for research community.

## Notes

### Competing Interest Statement

The authors have declared no competing interest.

### Funding Statement

Not Applicable

